# Longitudinal analysis of the gut microbiome in adolescent patients with anorexia nervosa: microbiome-related factors associated with clinical outcome

**DOI:** 10.1101/2023.07.18.23292809

**Authors:** Nadia Andrea Andreani, Arunabh Sharma, Brigitte Dahmen, Hannah E. Specht, Nina Mannig, Vanessa Ruan, Lara Keller, John F. Baines, Beate Herpertz-Dahlmann, Astrid Dempfle, Jochen Seitz

**Author notes:** These authors contributed equally to present study. This author serves as corresponding author: Nadia Andrea Andreani, Max Planck Institute for Evolutionary Biology, August-Thienemann-Str. 2, 24306 Plön, Germany Author ORCID: Nadia Andrea Andreani: 0000-0003-0330-9874 Arunabh Sharma: 0000-0001-7808-0942 Brigitte Dahmen Hannah E. Specht Nina Mannig Vanessa Ruan Lara Keller John F. Baines Beate Herpertz-Dahlmann: 0000-0001-8450-3323Astrid Dempfle: 0000-0002-2618-3920Jochen Seitz: 0000-0002-0110-7980.

## Abstract

There is mounting evidence regarding the role of gut microbiota in anorexia nervosa (AN). Previous studies have reported that patients with AN show dysbiosis compared to healthy controls (HCs); however, the underlying mechanisms are unclear, and data on influencing factors and longitudinal course of microbiome changes are scarce. Here, we present longitudinal data of 57 adolescent inpatients diagnosed with AN at up to nine time points (including a 1-year follow-up examination) and compare these to up to six time points in 34 HCs. 16S rRNA gene sequencing was used to investigate the microbiome composition of fecal samples, and data on food intake, weight change, hormonal recovery (leptin levels), and clinical outcomes were recorded. Differences in microbiome composition compared to HCs were greatest during acute starvation and in the low-weight group, while diminishing with weight gain and especially weight recovery at the 1-year follow-up. Illness duration and prior weight loss were strongly associated with microbiome composition at hospital admission, whereas microbial changes during treatment were associated with kilocalories consumed, weight gain, and hormonal recovery. The microbiome at admission was prognostic for hospital readmission, and a higher abundance of *Sutterella* was associated with a higher body weight at the 1-year follow-up. Identifying these clinically important factors further underlines the potential relevance of gut microbial changes and may help elucidate the underlying pathophysiology of gut-brain interactions in AN. The characterization of prognostically relevant taxa could be useful to stratify patients at admission and to potentially identify candidate taxa for future supplementation studies aimed at improving AN treatment.

## Introduction

Anorexia nervosa (AN) is the third most common chronic disease in adolescence and the deadliest of all psychiatric diseases, with a standardized mortality rate 5-10 times higher than that in healthy controls (Arcelus et al., 2011; Bulik et al., 2019; Carr et al., 2016). AN is characterized by insufficient energy intake, low body weight, body image distortion, and fear of gaining weight. However, the underlying pathophysiology is poorly understood. Treatment includes weight restoration and psychotherapy but often remains insufficient, and there is a high rate of relapse (Herpertz-Dahlmann et al., 2021).

The gut microbiome is increasingly recognized as an influencing factor for energy extraction from food and weight regulation, as well as having an influence on the brain and behavior via the gut-brain axis. Interest in the role of the microbiome in psychiatric diseases is on a steep rise (Hills et al., 2019; Santacroce et al., 2021; Wu et al., 2021).

Animal models of AN show intestinal dysbiosis (Breton et al., 2021; Trinh et al., 2021) and point to the potentially important role of gut microbes in the pathogenesis and course of AN. Offspring of gnotobiotic mice transplanted with AN patients’ stool showed reduced weight gain, as well as increased anxiety and obsessiveness, which are common comorbidities in AN (Hata et al., 2019). Moreover, directly transplanted “humanized” mice with AN patients’ stool showed lower weight gain than HC-transplanted animals when fed a calorie-reduced diet (Fan et al., 2023).

Patient studies during acute starvation have confirmed intestinal dysbiosis, albeit with heterogeneous results (Di Lodovico et al., 2021; Mack et al., 2016; Seitz et al., 2020). Very few longitudinal studies have reported residual changes after short-term weight restoration (Fouladi et al., 2022; Mack et al., 2016; Schulz et al., 2021). However, to our knowledge, no study has followed patients after hospital discharge or tracked them for longer than 6 months. Thus, it remains unclear whether the microbiome normalizes over a longer period after weight gain. Moreover, hospital food can act as a potential confounder when comparing the microbiome to healthy controls who eat at home. Furthermore, while studies have identified clinical factors to be associated with the microbiome in AN cross-sectionally (Borgo et al., 2017; Di Lodovico et al., 2021; Fan et al., 2023; Mack et al., 2016; Yuan et al., 2022), there are no reports of longitudinal associations, and only one study has attempted to predict the clinical course from microbiota alterations found at admission in AN (Schulz et al., 2021).

Here, we present the first longitudinal investigation, including data collected at the 1-year follow-up after admission, to investigate the degree of microbial normalization in individuals with AN classified as weight-recovered and on a home-based diet (thus making hospital food-related effects less likely). Up to nine time points were sampled to determine which clinical factors influence the microbiome in patients with AN, including illness duration, weight loss, and body weight at admission, and the potentially differential influence of nutritional, weight-related, and hormonal restitution during the treatment process. Furthermore, this longitudinal study allowed us to test whether microbiota can help predict weight development and relapse.

## Results

Fifty-six patients aged between 12 and 20 years and diagnosed with AN or atypical AN (one patient) according to DSM-5 were admitted to the specialized inpatients eating disorder unit at the Department for Child and Adolescent Psychiatry of the RWTH Aachen University Hospital. Stool samples and clinical data were collected at up to eight timepoints (T0-T7) during inpatient stay with one additional sampling at follow-up appointment (T8) 1 year after admission (Fig 1). After discharge, eight patients were re-admitted to the department within one year due to weight loss. At follow-up assessments, patients were classified as having low weight or still weight recovered based on the age- and sex-specific percentile of the Body Mass Index (BMI) based on the German Health Interview and Examination Survey for Children and Adolescents (KiGGS; Neuhauser et al., 2013). Specifically, individuals with a BMI lower than the 15^th^ percentile (P<15) were classified as low weight, while individuals with a BMI greater than or equal to the 15^th^ percentile (P≥15) were classified as weight-recovered (chosen with a safety margin of 5 percentile points towards the official definition of “underweight” at the 10^th^ BMI percentile). Inpatient treatment included weight rehabilitation with incremental increases in kilocalories (Fig 2A) and weight gain until achieving individually determined target weight, based on weight before the onset of the illness, hormonal recovery, and menstruation state (Fig 2B). Additionally, serum leptin concentration was measured at admission, discharge, and follow-up appointments to test for hormonal recovery (Fig 2C). Thirty-four age-matched healthy controls (HCs; aged between 14 and 19 years) were sampled at six time-matched time points (Fig 1). A summary of the clinical characteristics of the patients included in this study is presented in Tab 1.

**Figure 1:**
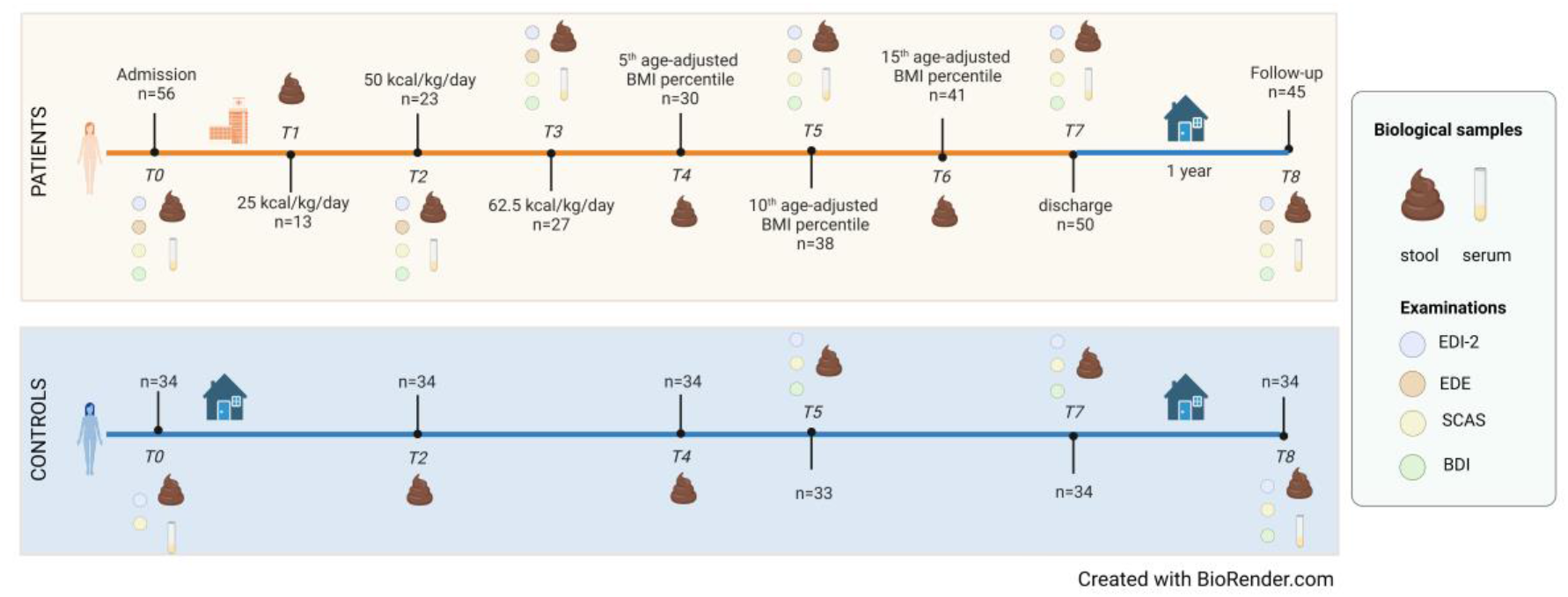
overview of the longitudinal study. This figure summarizes the samples used in this study, where n refers to the number of individuals included at each time point.

**Table 1:**
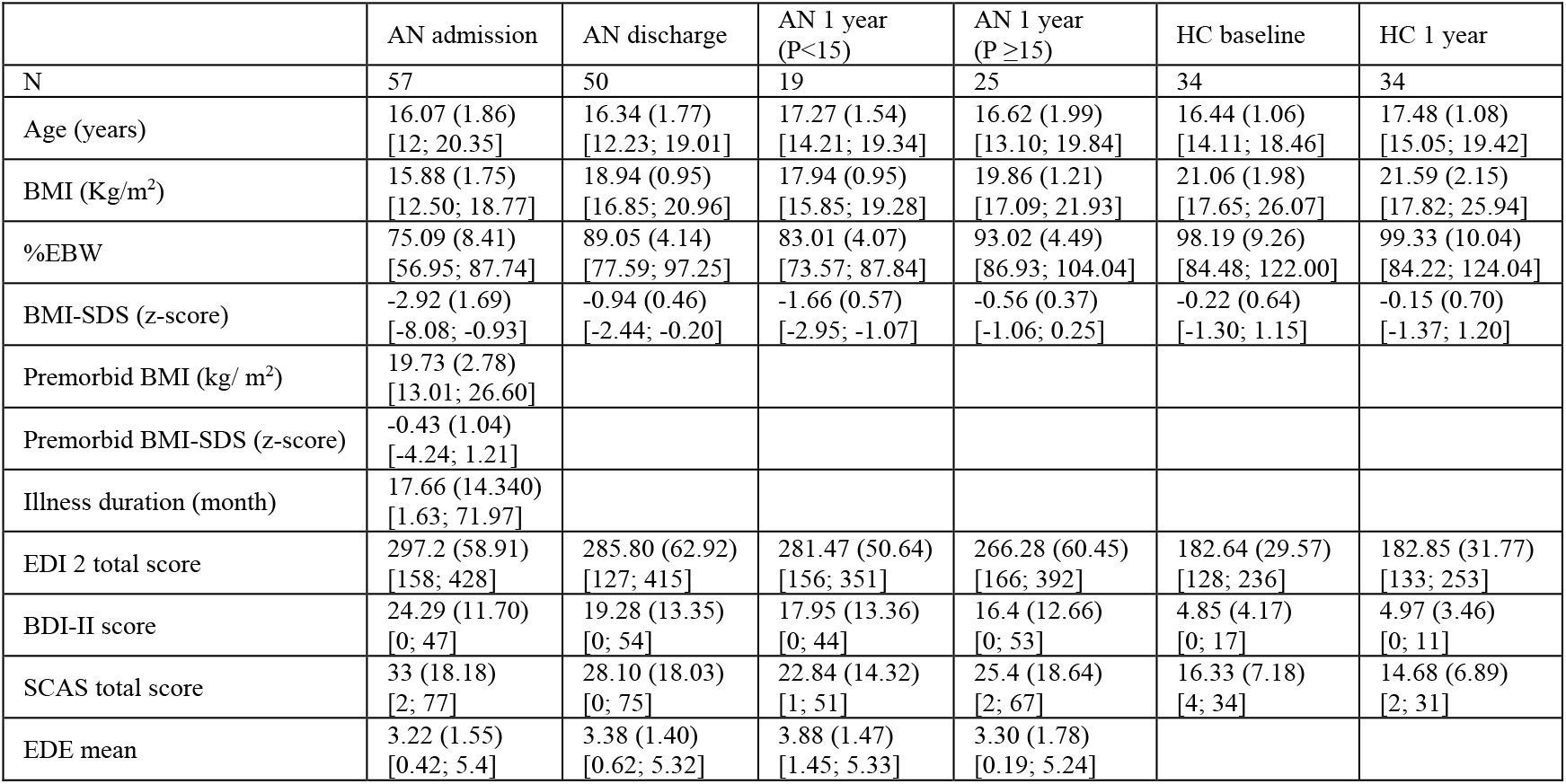
Clinical sample characteristics. Values are reported as the mean and standard deviation in brackets, and minimum and maximum values in square brackets. Abbreviations: AN, Anorexia Nervosa; BDI, Beck Depression Inventory; BMI-SDS, Body mass index-standard deviation score; EDE, Eating Disorder Examination; EDI, Eating Disorder Inventory; HC, Healthy Controls; SCAS, Spence Children’s Anxiety Scale; %EBW: Percent expected body weight.

**Figure 2:**
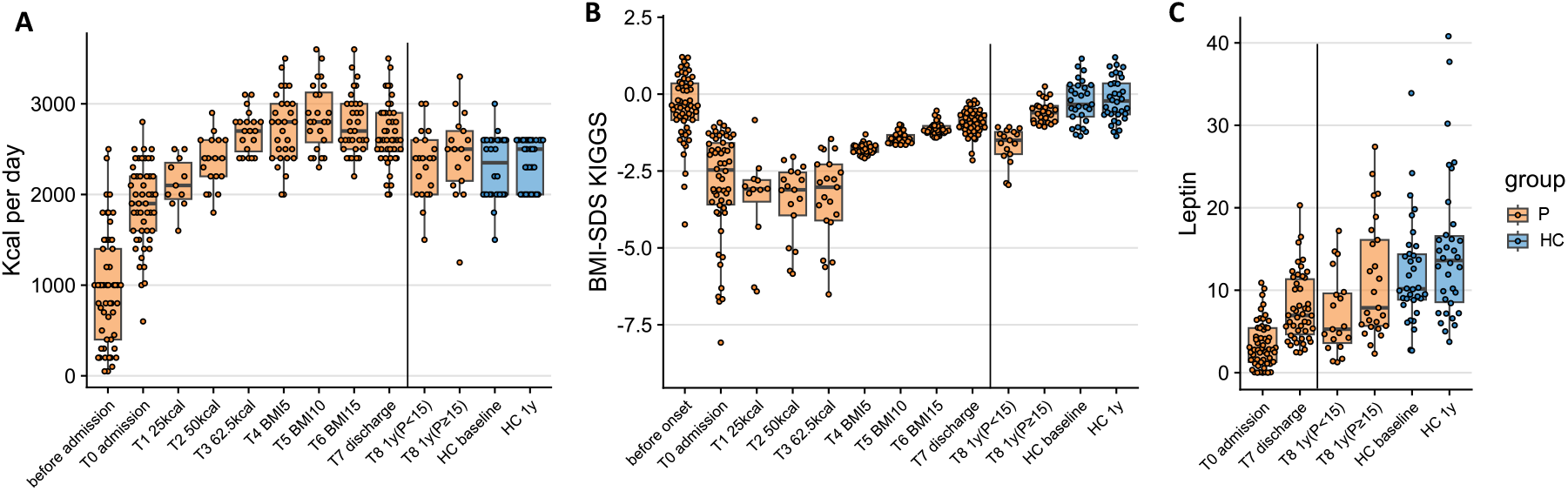
clinical information of the patients and age-matched healthy controls. The figure reports: **(A)** the amount of daily caloric intake of patients before admission, during inpatient treatment, and at follow-up appointment (1 year after admission), compared to HCs at baseline and at 1 year; **(B)** the BMI-SDS of patients before disease onset, during inpatient treatment, and at follow-up appointment (1 year after admission), compared to HCs at baseline and at 1 year; and **(C)** the serum concentration of leptin (ng/ml) of patients at admission, discharge, and at follow-up appointment (1 year after admission), compared to HCs at admission and at 1 year.

Inpatient treatment aimed at body weight rehabilitation as well as at improving eating behavior, as evidenced by the increase in BMI, percentage of Expected Body Weight (%EBW), and BMI-SDS, despite these measures being still different from the HC group (Tab 1). After 1 year, patients with a BMI at or above the 15^th^ percentile (classified as weight-recovered) had clinical characteristics that were more similar to the age-matched HC group (Tab 1 and Fig 2).

### Differences in gut microbiome between AN patients and healthy controls and longitudinal changes in the gut microbiome

A total of 22,848,660 16S rRNA sequence reads (63,645.29 ± 25,857.34 per sample, mean ± SD) were generated from the DNA extracted from 359 stool samples at nine time points, as specified in the Patients and Methods section and summarized in Tab S1_Sheet1.

After clustering with >97% similarity and rarefying 10,400 reads, 1,011 bacterial amplicon sequence variants (ASVs) were identified, which spanned 12 different phyla, 79 different families, and 212 different genera. Tab S1_Sheet2 shows the total ASV counts and taxonomic classification.

The gut microbiome of adolescent patients with AN was significantly different from that of HCs during acute starvation at admission (Fig 3A/B, Tab S1_Sheet3). The microbiome changed over the course of treatment but remained at least partly different from HCs at all time points, even in those patients who were classified as weight-recovered at 1-year follow-up [T8 1y(P≥15)]. Fig 3A shows the average relative abundances of 23 genera that were significantly different in pairwise comparisons of patients at admission, discharge, and follow-up assessment, as well as in comparison to HCs at admission and 1-year follow-up (Wilcoxon signed-rank, fdr corrected *p* < 0.05; see the following paragraphs and Tab S1 for details). Similarly, Fig 3B shows the shift in the microbiome composition in patients at all time points available between admission and follow-up, as seen in longitudinal multilevel partial least squares-discriminant analysis (mPLS-DA).

**Figure 3:**
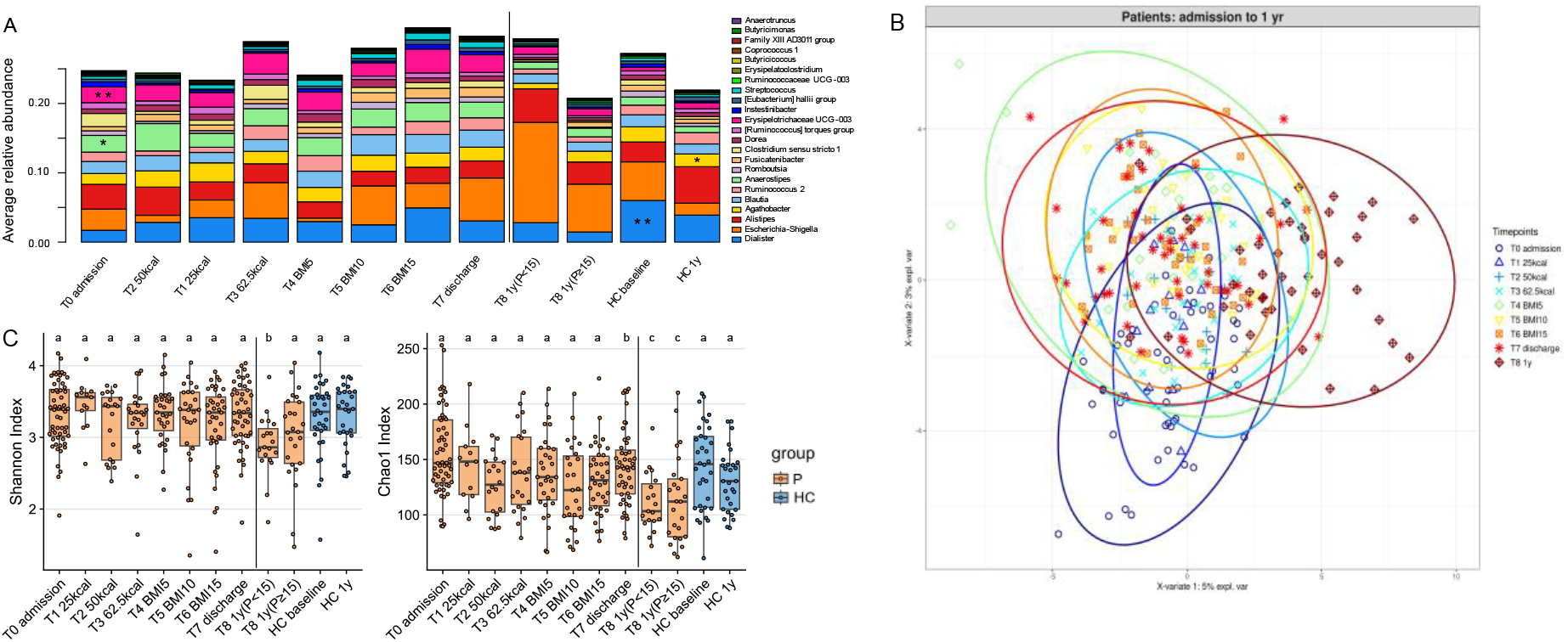
the microbiome of patients changes during inpatient treatment and differs from HCs. The figure reports: **(A)** the average relative abundance of 23 genera that were significantly differentially abundant in univariate pairwise comparisons of patients at different time points (T0, T7, and T8), and of healthy controls at baseline and 1 year. Stars at T0 admission report the main genera that have significantly higher abundance in fecal samples of patients at admission than in the respective HCs. For other pairwise comparisons, please refer to supplementary tables. **(B)** Multilevel PLS-DA of microbiome composition of patients at all time points; **(C)** Boxplots of alpha-diversity measures, Shannon Index (left), and Chao1 Index (right) of patients’ microbiome at different time points and of healthy controls. Groups depicted with the same letter show no significant differences when pairwise comparisons are performed.

Longitudinal changes in alpha-diversity using Shannon and Chao1 indices are displayed in Fig 3C. Both values were lowest in the two follow-up groups. Alpha-diversity indices of fecal samples of low-weight patients at the 1-year follow-up were significantly different compared to their values at admission and discharge time points and to HCs (Fig 3C, left, and Tab S1_Sheet3). Additionally, the Chao1 index showed a reduction during inpatient treatment, with significant differences when comparing patients at admission, discharge, and 1-year follow-up. Importantly, both values showed significant or trend-level reductions in weight-recovered patients after 1 year in comparison to HCs, hinting at a lack of complete microbiome recovery even in the weight-recovered subgroup (Fig 3C, right, and Tab S1_Sheet3).

To test the hypothesis that fecal microbiomes of patients with AN have a distinctive composition both cross-sectionally and longitudinally, we used a combination of univariate and multivariate approaches by comparing the microbiome of patients at different taxonomic levels (from phylum-to ASV-level) with the microbiome of the HC groups (Fig 4A, and Tab S1_Sheet4). PERMANOVA based on Bray-Curtis dissimilarities showed significant differences in overall microbiome composition between patients with AN at admission and HCs at baseline from family- to ASV-level (PERMANOVA *p*=0.0007 to 0.005, Fig 4A). PLS-DA showed that the genera *Legionella, Dialister, Ruminococcaceae UCG-003* and *Limnobacter* contributed most strongly to this differentiation, all of which were less abundant in AN (Fig 4B and Fig S1A). Univariate comparisons between the two groups in individual taxa at the family- to ASV-level identified differences in the genera *Erysipelotrichaceae UCG-003, Dialister, Family XIII group, Anaerostipes, Ruminococcaceae UCG-003, Anaerotruncus* and *Erysipelatoclostridium* (Mann-Whitney-U-test multiple comparisons fdr corrected *p=*0.0032 to 0.048, Tab S1_Sheet 5).

**Figure 4:**
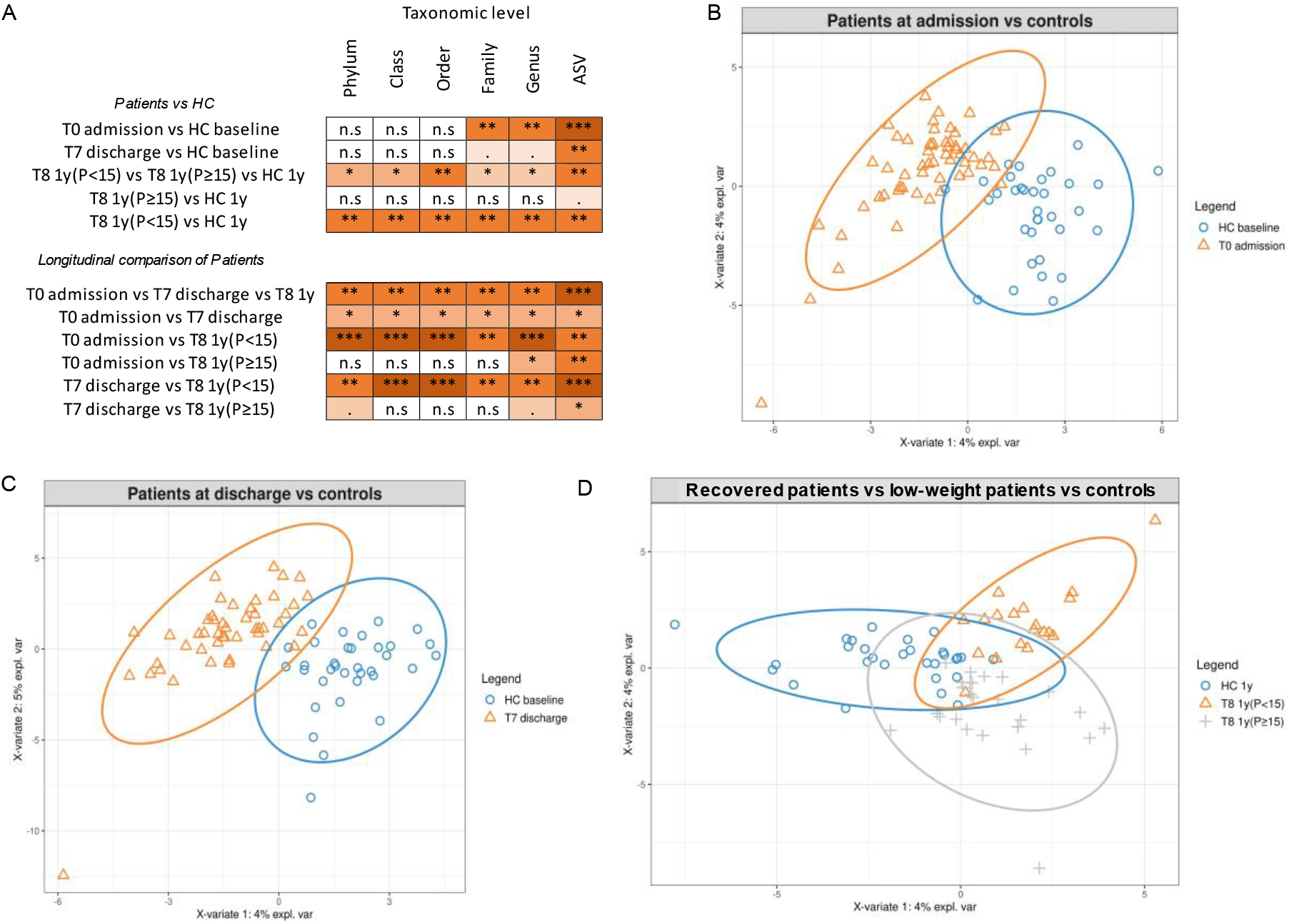
microbiome differences of patients at different timepoints and HC group. The figure reports: **(A)** a summary of the results of PERMANOVA test on Bray Curtis dissimilarities between patients and HCs, or between patients at different time points (n.s: p-value>0.05; *: p-value <0.01; **: p-value <0.001; ***: p-value <0.0001). Details are reported in Tab S1_Sheet4; **(B)** PLS-DA plots highlighting differences at the genus level between HCs at baseline and patients at admission, **(C)** at discharge, **(D)** and differences at the genus level between patients at the 1-year follow-up (divided into low-weight and recovered) and HCs at the 1-year follow-up.

At T7 discharge, PLS-DA and PERMANOVA based on Bray-Curtis dissimilarities showed a significant difference between AN and HC fecal microbiome only at the ASV-level (PERMANOVA *p*=0.007; Fig 4A, 4C, Tab S1_Sheet4, and Fig S1B), suggesting that inpatient treatment reduces the differences between these two groups. The Wilcoxon rank sum test identified five ASVs as being differentially abundant in patients at discharge compared with HCs. These were the ASV322 (uncl. *Erysipelotrichaceae UCG-003*), ASV524 (uncl. Lachnospiraceae), ASV578 and ASV798 (both uncl. *Ruminococcaceae UCG-005*) and ASV363 (uncl. *Dialister*) with a fdr corrected *p*=0.03 to 0.04 [Mann-Whitney-U-test, Tab S1_Sheet 6].

At T8_1-year follow-up, the microbiome of subgroups of patients with AN and HCs were again significantly different at all taxonomic levels (PERMANOVA *p*=0.005 to 0.03, Fig 4A, Tab S1_Sheet4), as also visible in the PLS-DA graph in Fig 4C and Fig S1C, which depict the main differences among low-weight, recovered patients, and the HC group. However, separately comparing low-weight patients [T8 1y(P<15)] with the HCs at 1 year showed comparable differences to admission at lower taxonomic levels (PERMANOVA *p*=0.001 to <0.001), with differences being recorded at the phylum-, class- and order-levels (Fig 4A, Fig S1D, and Tab S1_Sheet4). On the other hand, differences between recovered AN patients [T8 1y(P≥15)] and HCs were much smaller (PERMANOVA *p*=0.063) at the ASV-level when performing a PERMANOVA on Bray-Curtis dissimilarity distances (Fig 4A, Fig S1E and Tab S1_Sheet4). Univariate comparisons of low-weight patients with HCs revealed some major differences at all taxonomic levels (from phylum- to genus-level), with corrected *p*-values between 0.008 and 0.04 (Mann-Whitney-U-test, Tab S1_Sheet 7). The genera *[Eubacterium] hallii* group and *Agathobacter* were significantly more abundant in HCs than in low-weight patients. Interestingly, these differences were not observed when comparing recovered individuals with the HC group, with only one unique and rare taxon (family Desulfovibrionaceae) significantly more abundant in recovered patients (Mann-Whitney-U-test fdr corrected *p*=0.02; Tab S1_Sheet 8).

To understand the extent to which these changes are due to inpatient treatment, weight gain, and remission and to study the potentially confounding role of different foods in the hospital and at home, we performed pairwise investigations by applying multivariate and univariate approaches, comparing patients at admission, at discharge, and at the 1-year follow-up appointment. Inpatient treatment was associated with changes at all taxonomic levels (change between admission and discharge; PERMANOVA *p*=0.003 to 0.01). However, univariate analysis identified the genus *Fusicatenibacter* as a unique taxon that was significantly more abundant at discharge (Wilcoxon signed-rank test fdr corrected *p*=0.027; Tab S1_Sheet 9). Interestingly, among low-weight patients, the overall microbiome composition was significantly different between admission and 1-year follow-up, with PERMANOVA on Bray-Curtis dissimilarity showing significant *p*-values at all levels, potentially due at least in part to hospital food being consumed at admission vs. home food at follow-up (PERMANOVA *p*=0.0005 to 0.001; Fig 4A, and Tab S1_Sheet4). Specifically, low-weight patients showed a significant reduction in the relative abundance of the genera *Anaerostipes*, *Clostridium sensu stricto 1* and *Romboutsia* compared to the microbiome composition of these patients at admission (Wilcoxon signed-rank test corrected *p*-value= 0.02, Tab S1_Sheet 10). On the other hand, individuals who recovered at follow-up showed a surprisingly higher similarity between admission and follow-up, with significant *p*-values only at the genus- and ASV-level (Fig 4A and Tab S1_Sheet4).

Similarly, low-weight patients had a distinct microbiome at follow-up when compared to the assessment at discharge (PERMANOVA *p*=0.006 to 0.001, Fig 4A, and Tab S1_Sheet4), while these differences were less marked when comparing recovered patients between follow-up and discharge, with a significant *p*-value of the PERMANOVA only at the ASV-level (PERMANOVA *p* = 0.02). In low-weight patients, we observed 4 times higher abundance of the genus *Escherichia-Shigella* (Wilcoxon signed-rank test fdr corrected *p* = 0.04) and a 2 times higher abundance of *Alistipes* (Wilcoxon signed-rank test fdr corrected *p* = 0.03) between follow-up and discharge.

### Clinical variables associated with microbiome composition

To investigate which clinical variables were associated with the overall microbiome composition, we applied PERMANOVA analysis to the Bray-Curtis dissimilarity index. Analysis at admission revealed that illness duration (phylum-family level, PERMANOVA *p*=0.011 to 0.022) and the amount of weight loss (class-genera level, PERMANOVA *p*=0.030 to 0.047, Fig 5 and Tab S1_Sheet 13) were significantly associated with microbiome composition. In contrast, BMI-SDS (PERMANOVA *p*=0.14 to 0.24), Kcal at admission (PERMANOVA *p*=0.42 to 0.92), and leptin concentration (PERMANOVA *p*=0.26 to 0.84) displayed little variability in the acute starvation phase and did not show a significant association with microbiome composition at this time point.

**Figure 5:**
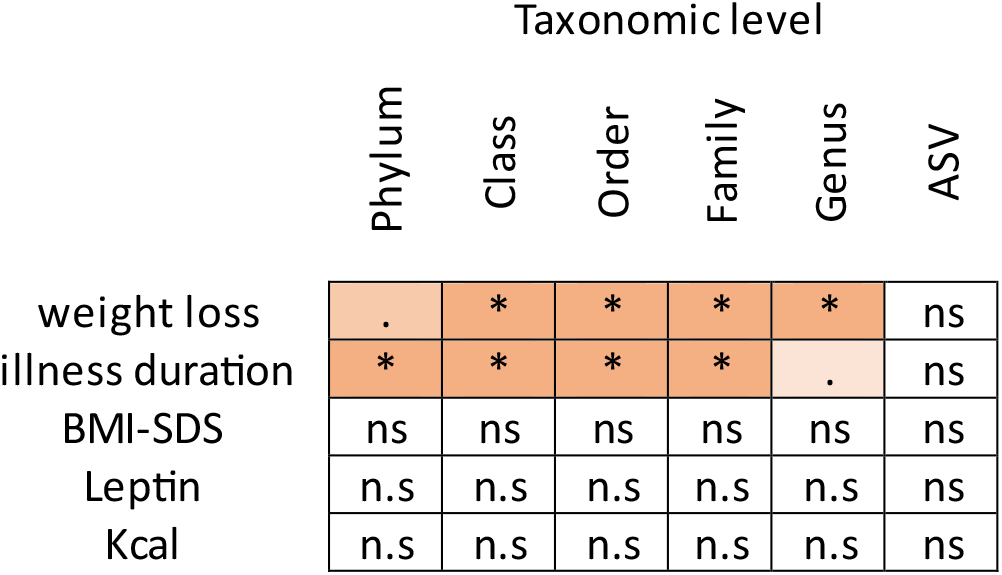
clinical variables that are associated with microbiome composition. n.s: p-value>0.05; *: p-value <0.01; **: p-value <0.001; ***: p-value <0.0001

Next, we performed a longitudinal PERMANOVA analysis, including genus data from all nine time points. After correcting for laxative use, we found a significant association between the gut microbiome and the amount of ingested Kcals (PERMANOVA *p*=0.003) and BMI-SDS (PERMANOVA *p*=0.005). Even though these two variables are highly correlated, they both showed independent contributions when combined in one model (PERMANOVA *p*=0.03 for BMI-SDS; *p*=0.02 for Kcals, respectively). Leptin concentration was measured at admission, discharge, and 1-year follow-up, and showed a significant association with the microbiome when analyzed alone (PERMANOVA *p*=0.02). When analyzed together, these three potentially influencing factors (Kcal, BMI-SDS, and leptin) were strongly intercorrelated, so that only BMI-SDS and leptin at the trend-level showed independent contributions (PERMANOVA *p*=0.023 to 0.054).

### Prediction of hospital-readmission and BMI-SDS at 1-year follow-up based on baseline microbiome data

PERMANOVA analysis of the whole microbiome community at admission revealed a significant association with hospital readmission until the 1-year follow-up when considering the data at admission at the ASV-level (*p*=0.04). The genera significantly associated with hospital readmission were *Ruminiclostridium 5* and *Intestinibacter* (*p*=0.006 and 0.03, respectively); ASVs showing significant association with hospital readmission until the 1-year follow-up were ASV600 and ASV95 (*p*=0.0004 and *p*=0.02, both uncl. *Subdoligranulum*), ASV666, and ASV83 (*p*=0.003 and *p*=0.008, both uncl. *Lachnospiraceae)*, ASV 238 (*p*=0.01, uncl. *Clostridium sensu stricto*), ASV19 (*p*=0.03, uncl. *Ruminiclostridium*) and ASV873 (*p*=0.03, uncl. *Intestinibacter*). There was no significant association between BMI-SDS at T8_1 year follow-up and overall microbiome composition at baseline (all *p*>0.2).

Linear model analysis related to the abundance of specific taxa at baseline to BMI-SDS at 1-year follow-up, while correcting for laxative use, illness duration, weight loss, and BMI-SDS at admission, identified four genera (*Sutterella, Parasutterella, Lachnospiraceae FCS020 group,* and *Clostridium sensu stricto, p*=0.008 to 0.040; Fig 6, Tab S1_Sheet14) and four ASVs (uncl. *Clostridium sensu stricto*, uncl. Bacteroides, uncl. *Alistipes*, uncl. *Parasutterella*, *p*=0.002 to 0.01, Fig 6, Tab S1_Sheet14) that were associated with the BMI-SDS at 1 year follow-up.

**Figure 6:**
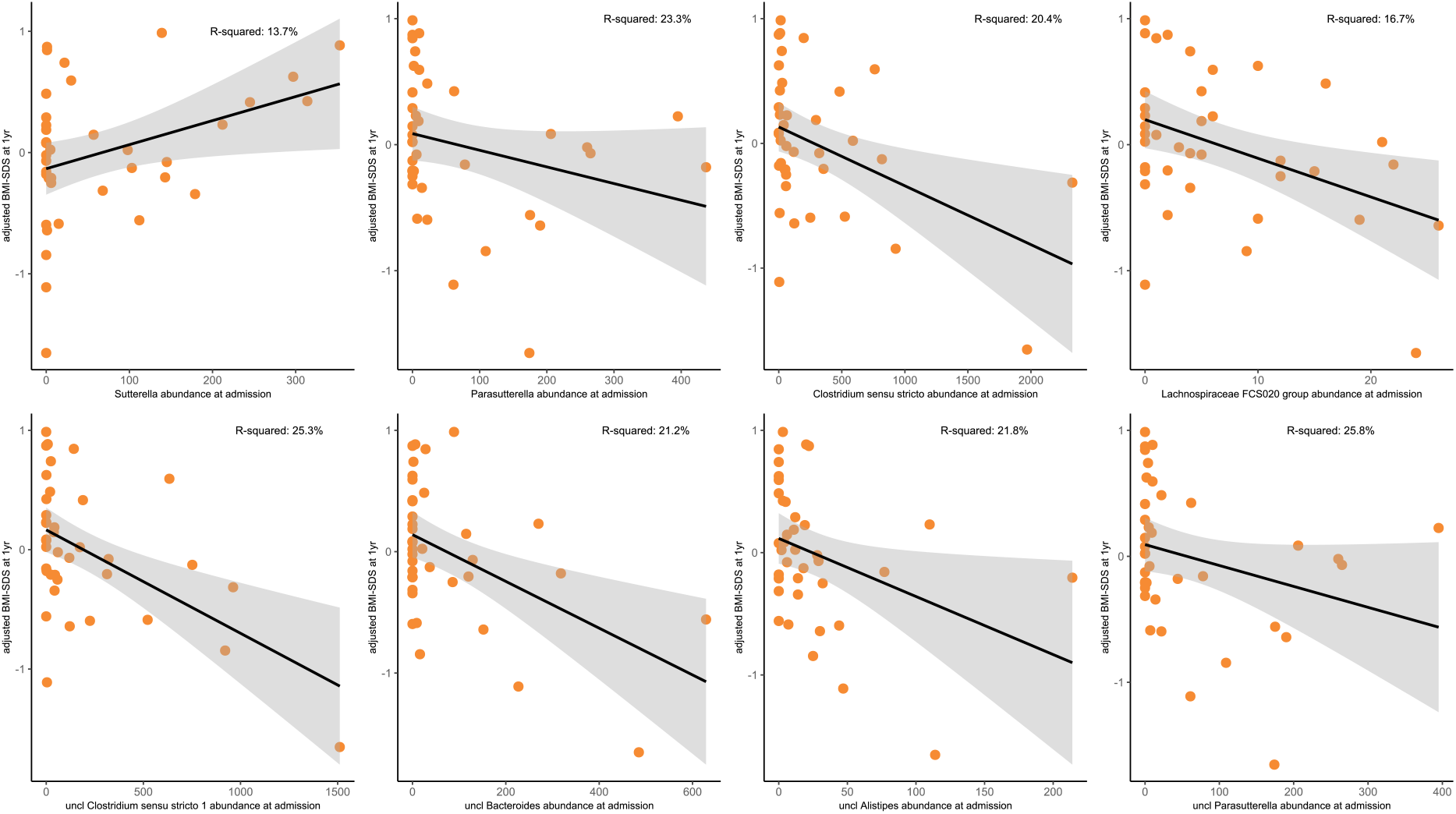
taxa predicting BMI-SDS at 1-year follow-up. Association of specific taxa at admission (T0) with BMI-SDS at 1-year follow-up, adjusted for weight loss (before admission), duration of illness, BMI-SDS at admission, and laxative use (see Patients and Methods for details on statistical models).

## Discussion

Our study presents the first longitudinal investigation of fecal microbial changes in patients with AN compared with age-matched healthy controls, including a 1-year follow-up. Our results show that dysbiosis in acutely ill patients with AN is improved during inpatient treatment and long-term weight recovery. The remaining microbiome alterations in the weight-recovered subgroup after 1 year compared to HCs were small but remained significant. The differences found in the low-weight group indicate that the disease itself is the main driver of these microbiome differences and not the consequence of the consumption of hospital vs. home food. The up to nine assessment points sampled in this study allowed us to further elucidate the clinical factors associated with these longitudinal microbial changes. We found that illness duration and the amount of weight loss prior to admission were important for microbiome composition at admission, in line with an extensive body of research showing their clinical relevance as markers of disease severity. We also found that the kilocalories consumed, ensuing weight gain, and hormonal restitution were all clearly related to longitudinal microbiome composition changes during the treatment process. These results indicate that starvation is a major driving force for changes in the microbiome. These results further support previous clinical findings regarding the importance of nutritional restitution and reaching a sufficiently high target weight. Finally, our longitudinal follow-up showed that the composition of microbiota at admission can help predict relapse and that individual taxa were associated with increased or decreased BMI-SDS at the 1-year follow-up. Importantly, a higher abundance of *Sutterella* is indicative of a positive clinical outcome and thus qualifies as a potential probiotic target or supplement for future animal and human studies. These findings are in line with a potentially important or contributing role of the gut microbiome on clinical outcomes, at least as a factor in maintaining the disease, as suggested by animal studies.

### Altered microbiome characteristics in patients with AN

Our data report a reduction in Shannon’s and Chao 1 indices at 1-year follow-up, which were slightly more marked in low-weight patients. We did not observe any significant change in alpha-diversity measures during inpatient treatment, as found by Kleiman et al. (2015); however, the literature is heterogeneous regarding this point (Dhopatkar et al., 2023; Di Lodovico et al., 2021; Garcia & Gutierrez, 2023). The reduction in alpha-diversity after returning to a home environment could be associated with a change in diet, although this does not explain the newly appearing differences in the comparison with healthy controls. Most likely, different underlying mechanisms overlap in influencing alpha-diversity at this point in time, including remaining differences in food choice, body weight, hormonal status, and exercise, even in recovered patients with AN (Castro et al., 2021; Hübel et al., 2019).

Although the gut microbiome in patients with AN remained different from that in HCs at all time points, the differences diminished with weight recovery and over time. At admission, there were significant multivariate and univariate differences at different taxonomic levels, whereas after short-term weight recovery at discharge, only ASVs showed significant differences. This is in line with previous studies suggesting that the composition of the gut microbiome shifts during inpatient weight gain treatment (Fouladi et al., 2022; Kleiman et al., 2015; Mack et al., 2016; Schulz et al., 2021). The fact that patients with AN have a distinct microbiome when compared to healthy individuals is in line with several studies that have focused on the fecal microbiome in adults (Borgo et al., 2017; Fan et al., 2023; Monteleone et al., 2021), or the combination of both adolescents and adults combined (Mack et al., 2016; Yuan et al., 2022).

One added value of our study was the presence of a long-term follow-up investigation 1 year after admission. Interestingly, the microbiome of weight-recovered individuals (i.e., those who had a BMI at least greater than or equal to the 15^th^ percentile) showed only small differences from that of the HCs (univariate differences in Desulfovibrionaceae family, a trend in the multivariate analysis and significant alpha-diversity differences). Importantly, as the target weight was determined at an individual level, many patients had a BMI higher than the 15^th^ percentile (mean 29^th^ percentile, range 15^th^-60^th^ percentile) considered necessary for their personal recovery. These results support the hypothesis of continuous recovery of the gut microbiome with adequate weight increase and/or time. However, a complete recovery cannot yet be concluded at the 1-year follow-up in weight-recovered patients. Further research is necessary to address this phenomenon, especially as the remaining differences in nutritional uptake and the remaining lower body weight compared with HCs are common in this subgroup, as noted above. Interestingly, the low-weight subgroup showed dramatic differences when compared to controls again at the 1-year follow-up, ruling out a hospital vs. home food artifact in previous comparisons of acutely ill (hospitalized) patients with AN and HCs at home. This further underlines the importance of sustaining a healthy weight and, also for gut microbial normalization.

At admission, *Dialister* was significantly less abundant in patients than in age-matched HCs (as also reported in Garcia-Gil et al., 2022 and Yuan et al., 2022); interestingly, this genus increased in abundance during inpatient treatment and was not significantly differentially abundant in any other pairwise comparisons. A large cohort study found a reduced abundance of this genus in people diagnosed with depression and major depressive disorder, suggesting a role for this taxon in contributing to the psychological signs associated with AN (Cheung et al., 2019; Valles-Colomer et al., 2019). Another notable increase in *Escherichia* abundance was observed between discharge and follow-up appointments in patients with insufficient weight recovery. *E. coli* is a well-known producer of caseinolytic protease B (ClpB), a protein involved in appetite regulation (Breton et al., 2021; Tennoune et al., 2014), and associated with anxiety severity (Mason et al., 2020) and AN (Yuan et al., 2022). Remarkably, this genus appears to be differentially abundant only in low-weight patients, suggesting its involvement in the chronicity of the disease. Similarly, the genus *Alistipes* was significantly more abundant in these patients (on average twice as abundant as that at discharge). A member of *Alistipes* (*Alistipes ihumii*) was isolated for the first time in 2014 from a patient with AN (Pfleiderer et al., 2014). Members of this genus are involved in anxiety and depression, as they have the potential to degrade tryptophan, the precursor of serotonin (Garcia-Gil et al., 2022; Kaur et al., 2019; Parker et al., 2020), and have been reported to be increased in patients with AN when compared to HCs (Di Lodovico et al., 2021; Prochazkova et al., 2021; Schulz et al., 2021). Overrepresented taxa in low-weight patients also include the genus *Anaerostipes*, which was found to be increased in anorexia nervosa, bulimia nervosa, and depression (Leyrolle et al., 2021). This is in line with previous research showing an increase in mucin-degrading taxa, such as *Escherichia-Shigella, Alistipes, Anaerostipes* (Di Lodovico et al., 2021; Glover et al., 2022) and a reduction in butyrate producers, and an increase in carbohydrate (fiber)-degrading taxa (Fouladi et al., 2022). In our longitudinal study, we failed to detect any major perturbation in a butyrate-producing taxon (Singh et al., 2023).

Low-weight patients show a reduction in *Ruminococcus*, a well-known beneficial carbohydrate-fermenter commensal, whose abundance has increased in different studies as a consequence of increased fiber intake (Kleiman et al., 2015; Mack et al., 2016; Ruusunen et al., 2019). Underrepresented genera in the AN microbiome of low-weight patients include *Agathobacter* (also as reported by (Prochazkova et al., 2021; Yuan et al., 2022)) and *Romboustia* (as in (Yuan et al., 2022)). Interestingly, some of the taxa that were overrepresented in patients at admission and in low-weight patients were the same and were reported to be higher in stunted children: Family XIII AD3011 group, uncl. Erysipelotrichaceae, and uncl. Ruminococcaceae (Golloso-Gubat et al., 2020).

After a 1-year follow-up, the family Desulfovibrionaceae remained altered even in the weight-recovered subgroup, with a relative abundance five times higher in recovered individuals than in HCs. These bacteria were shown to be associated with binge eating disorders in obese individuals (Navarro-Tapia et al., 2021), although they represent a rare taxon (less than 0.20% of the total microbiome) in our study, and it is thus difficult to disentangle their role in the gut-brain axis.

Importantly, the diminishing difference between HCs and recovered AN patients was immediately offset in the case of a repeated weight loss. Low-weight patients showed renewed dysbiosis at all taxonomic levels of similar magnitude as at admission. This emphasizes the importance of maintaining a healthy target weight for regaining a healthy gut microbiome.

### Clinical factors associated with gut microbiome

Interestingly, our analysis at admission showed that illness duration and weight loss were associated with alterations in the overall composition and structure of bacterial communities within the group of adolescent patients with AN. To our knowledge, this is the first time that these two variables have been linked to the microbiome at admission. This is potentially explained by the difference in age compared with adult patients, which is typically accompanied by a difference in illness duration. Studying adolescents might have the advantage of having a higher percentage of first-time ill patients with fewer compensatory or treatment-related factors, allowing for a less confounded study of the original underlying pathophysiology. Interestingly, the absolute low BMI-SDS did not reach significance cross-sectionally, which agrees with previous reports (Fan et al., 2023; Mack et al., 2016). However, Di Lodovico (Di Lodovico et al., 2021) identified a correlation between *Roseburia* abundance and BMI. Borgo et al. (Borgo et al., 2017) highlighted a negative correlation of *Bacteroides* in adult samples, while Yuan et al. (Yuan et al., 2022) found *Subdoligranulum* was positively and *Bacteroides* was negatively associated with BMI in a mixed sample of adolescents and adults. This might be explained by differences in the statistical approach (PERMANOVA vs. univariate single taxa analyses) or by differences in age and illness duration.

Our longitudinal PERMANOVA analysis including genus-level data from all nine time points (adjusted for laxative use) helped address the question of which clinical parameters and physiological changes during treatment were related to the changing gut microbiome. Indeed, we found that all three of our hypothesized factors (kilocalories consumed, achieved weight gain, and hormonal recovery) were strongly related to changes in microbiome composition. This is consistent with the previous clinical literature, which primarily underscores the influence of nutrition (David et al., 2014). It is interesting and important to demonstrate that it is not purely nutritional rehabilitation that drives changes in the microbiome. Also, body weight recovery was an important factor, potentially because of its link to an increase in fat mass, normalization of metabolism and leaving behind the “emergency-state” of semi-starvation with all its other metabolic counter-regulations to conserve energy. Finally, hormonal restitution, studied using leptin concentration in the serum, showed an individually and independently significant contribution. Leptin is an anorexigenic hormone secreted by fat cells and is known to be severely reduced in acutely ill patients with AN and to recover with weight gain. It has numerous effects on metabolism, and its accommodation to starvation and leptin receptors is found virtually throughout the human body (Hebebrand et al., 2022). Interestingly, it is known to be both affected by (Yao et al., 2020) and to affect gut bacteria (Neuman et al., 2015), and has recently been shown to be very promising as an experimental treatment in chronic patients with AN (Gradl-Dietsch et al., 2023; Milos et al., 2020).

Taken together, these findings are important for understanding the underlying microbiome-gut-host interactions during AN. By identifying clinically relevant factors associated with microbiome changes, they added further validity to the relevance of the microbiome being linked to the severity and course of the disease.

### Prognostic relevance of baseline microbiome for clinical outcome

After showing how clinical factors are associated with the microbiome in patients with AN, we also investigated the potential influence of the microbiome on the course of illness. This is the first study to show that microbiome composition at admission is prognostically relevant for hospital readmission within the first year. This shows the potential of microbiome analyses to help clinicians in the prognosis of the clinical course and potentially divert more intense resources to those most at risk. Interestingly, for most of the genera and taxa identified, a higher abundance was associated with a negative course (*Parasutterella, Clostridium sensu stricto; Lachnospiraceae FCS020 genera* and *uncultured Alistipes*). Thus, the high abundance of these taxa should be regarded as a risk factor. Interestingly, a recent cross-sectional study on obesity and type 1 diabetes reported the role of the genus *Parassuterella* in the stimulation of the biosynthetic pathways of fatty acids, suggesting a role in body weight gain. Moreover, *Parassuterella* was significantly reduced during weight loss interventions (Henneke et al., 2022). *Clostridium sensu stricto 1* (also known as *Clostridium* cluster 1) levels were 22 times higher at admission than at follow-up in low-weight patients. *Clostridium sensu stricto* 1 is a well-known mucin degrader that has been associated with AN, as this genus can induce a leaky gut (Di Lodovico et al., 2021; Kleiman et al., 2015; Mack et al., 2016). *Alistipes* is known to be involved in depression, which highlights the multifactorial nature of AN (Parker et al., 2020). Members of the family *Lachnospiracea* are known to degrade carbohydrates to produce butyric acid and other SCFAs, which could help reduce inflammation (Yuan et al., 2022). Decreased *Lachnospiraceae* is a well-accepted marker of inflammation in several inflammatory disorders (Geirnaert et al., 2017; Maukonen & Ouwehand, 2022). Patients with AN are thought to have chronic low-grade inflammation of unknown origin (meta-analysis by (Dalton et al., 2018)); however, recent findings are more diverse and show variable dysregulation of the immunologic state (Keeler et al., 2022; Schulz et al., 2021). AN is also associated with an increased rate of autoimmune diseases, especially Crohn’s disease and celiac disease (Hedman et al., 2019; Raevuori et al., 2014). Therefore, the immunomodulatory effects of these taxa might be important for their mechanistic role in AN outcomes.

In contrast, a higher *Sutterella* abundance at admission was associated with a positive outcome of increased body weight after one year. *Sutterella* are commensals associated with a positive pro-inflammatory status (Hiippala et al., 2016). They are known to modulate inflammatory processes; they decrease in multiple sclerosis and increase again after interferon therapy (Giri & Mangalam, 2019). In a mouse obesity study, *Sutterella* was not found in high-fat diet-fed animals and only appeared after the introduction of prebiotic treatment, associated with improved health (Everard et al., 2014). Due to the association of *Sutterella* with a positive clinical course in our study, they might also be interesting candidates for probiotic supplementation in Activity Based Anorexia (ABA)-animal models and patient studies.

These results complement our findings regarding clinical measures, such as illness duration or weight loss, being intertwined with the gut microbiome at admission. One possible interpretation is that the microbiome is initially influenced by food reduction and ensuing semi-starvation, fat mass reduction, and hormonal changes. Gut microbial changes could then exert a causal, upholding influence favoring the maintenance of the disease, as transplantation studies of stool of patients with ongoing AN into germ-free mice have shown reduced weight gain as well as brain and behavioral changes in animals similar to those in AN (Fan et al., 2023; Hata et al., 2019). Whether the gut microbiome plays a role in the initial occurrence of AN remains unclear and should be the target of future investigations.

Although this is one of the largest studies of patients with AN analyzing the microbiome, larger samples are still required regarding the sheer number of taxa involved and their potential interactions with each other and with the host. Furthermore, grouping patients at one year of admission together only on the basis of BMI is always somewhat artificial and demonstrates the dire need for a more stringent definition of recovery from an eating disorder including, for example, disordered eating behavior. Lastly, while longitudinal studies like ours present a major advancement compared to purely cross-sectional studies, they still only allow limited insight into causality and need to be supported by well-controlled interventional studies.

Taken together, we showed reduced, yet ongoing alterations in the gut microbiome after a 1-year follow-up, even in weight-recovered patients, and identified an important relationship between illness duration and weight loss with microbiome composition at admission and provided evidence that kilocalories, body weight, and hormonal recovery are all associated with the changing microbiome during treatment and weight gain. The microbiome at admission has prognostic value for the course and outcome of the disease. Our results are consistent with the hypothesis that changes in the body environment following semi-starvation influence the composition of the gut microbiome. Together with transplantation data, our study further supports the potential causal role of certain microbes, at least as maintaining factors prolonging the disease. The role of the microbiome in the etiology of the initial phase of disease warrants further investigation. Identifying taxa whose abundances are prognostic for the clinical course could help stratify patients at admission and increase therapy intensity where most needed, whereas *Sutterella* could potentially yield promising microbiome-targeted therapies as future additions to existing AN treatment.

## Patients and methods

### Recruitment

Sixty-four female adolescents (aged between 12 and 20, mean16 years) diagnosed with typical or atypical AN according to the DSM-5 were recruited at the Department for Child and Adolescent Psychiatry of the RWTH Aachen University Hospital and enrolled between December 2016 and January 2020. Seven patients dropped out of the study, leaving 57 patients for the analysis. The inclusion criteria were the same as those previously published before (Schulz et al., 2021) with minor modifications: diagnosis of AN according to DSM 5, female sex, and age between 12 and 20 years. The exclusion criteria were as follows: use of antibiotics or probiotics within four weeks before enrollment, IQ<85, insufficient knowledge of the German language, severe other mental disorders, and severe gastrointestinal or metabolic illnesses such as celiac disease or diabetes mellitus. Stool samples collected within four weeks of oral or intravenous antibiotic treatment were excluded from the analysis. Admission and discharge data for a subset of the current study (20 patients with AN and 20 HCs) have been published previously (Schulz et al., 2021).

Additionally, 34 age-matched female HCs with normal body weight (>20th and <80th age adjusted percentile of body mass index [BMI-SDS]) were enrolled using newspaper advertisements. The same exclusion criteria as above were applied to HCs, in addition to any current psychiatric illness or any lifetime eating disorder. All participants and their legal guardians provided written informed consent prior to enrollment. Consent was obtained from the ethics committee of the RWTH Aachen University Hospital for this study, and the study was conducted in accordance with the Declaration of Helsinki.

Clinical data included height and body weight after an overnight fast at admission and discharge, as well as weight and height prior to disease onset, weight loss prior to admission, and illness duration. For all time points, BMI as well as age- and sex-specific BMI percentiles and BMI-SDS were calculated based on German reference data from the KiGGS study (Neuhauser et al., 2013). Any medication used at admission was noted and sorted into the following groups for use as a binary covariate: laxatives, antibiotics, antidepressants, gastrointestinal medications, and others.

### Assessment timepoints

Up to nine time points were chosen for sampling (Fig 1): T0 (admission), T1 (corresponding to a diet of 25 Kcal/kg/day), T2 (corresponding to a diet of 50 Kcal/kg/day), T3 (corresponding to a diet of 62.5 Kcal/kg/day), T4 (corresponding to a weight gain up to the 5^th^ age-adjusted BMI percentile), T5 (corresponding to a weight gain up to the 10^th^ age-adjusted BMI percentile), T6 (corresponding to a weight gain up to the 15^th^ age-adjusted BMI percentile), T7 (discharge), and T8 (1-year follow-up appointment, one year after admission). Based on the clinical course, some patients reached more than one timepoint at a time and had fewer sampling time points. Thirty-four HCs’ samples were collected at six time points (T0, T2, T4, T5, T7, and T8; Fig 1).

### Questionnaires and interviews

Each participant in the study completed three questionnaires at admission, T2, T4, T5, discharge, and 1-year follow-up appointment: The Eating Disorder Inventory 2 (EDI-2; (Garner, 1991)), Beck Depression Inventory-II (BDI-II; (Kühner et al., 2007)) and Spence Children’s Anxiety Scale (SCAS; (Spence, 1998)). Patients also underwent semi-structured EDE (Eating Disorder Examination, first German edition 2016; (Cooper & Fairburn, 1987; Luce & Crowther, 1999)) at admission, discharge, and 1-year follow-up.

### Fecal sample collection and DNA extraction

Fecal samples were collected as previously described (Schulz et al., 2021). Healthy volunteers collected their stool at home using the same procedure and brought or sent the samples to the clinic to be frozen at −80°C until further use. DNA extraction from stool samples was performed using the DNeasy Power Soil Kit (Qiagen), following the manufacturer’s instructions.

### 16S rRNA gene sequencing and processing

The V1-V2 region of the 16S rRNA gene was amplified with primers 27F and 338R using dual barcoding. During demultiplexing, no mismatches were allowed in the barcode (Casava, Illumina). QIIME2 (v2019.10) was used to process and analyze the sequence data (Bolyen et al., 2018). Paired end sequences were denoised with ‘dada2’ (Callahan et al., 2016) using default parameters, unless stated: reads were truncated at the first base where the quality score dropped below Q=3, the maximum number of mismatches in the overlap region was 2, and the minimum length of reads after truncation was 250 bp. Merged sequences were clustered into amplicon sequences variants (ASVs) using ‘vsearch’ with an identity of 0.97 (Rognes et al., 2016). Bacterial ASVs were annotated using the q2-feature-classifier plugin (Bokulich et al., 2018). The sequences were rarefied at 10,400 reads per sample.

### Statistical analysis

All statistical analyses were performed using the R software (v. 4.1.1). Alpha-diversity within samples (represented by the Shannon and Chao1 indices) was determined by applying the estimate and diversity functions in “vegan” package (Oksanen & et al., 2022), while microbial dissimilarities between samples (beta-diversity defined by Bray-Curtis dissimilarity) were estimated at all taxonomic levels (phylum-to ASV-levels). Different time points of the HCs were used in our analyses, and the first available time point (baseline T0) was used to compare the microbiome compositions of HCs with those of patients with AN at admission (T0) and discharge (T7). The last available time point for each HCs (usually after one year, T8) was used for follow-up (T8) comparisons. We defined a ‘core’ microbiome as taxa that were present in at least 50% of individuals.

### Analysis of gut microbiome in AN patients and healthy controls and longitudinal changes in the gut microbiome

We used multivariate and univariate methods to compare the microbiomes of patients at various taxonomic levels (from phylum- to ASV-level) with a) the microbiomes of the HC group and b) those between different visits. To examine the differences in the overall microbiome composition between AN patients and the HC group at admission (T0), discharge (T7), and follow-up (T8), we used permutational multivariate analysis of variance (PERMANOVA; Anderson, 2017) implemented in the R package “vegan” (Oksanen et al., 2022) at all taxonomic levels. In all the PERMANOVA models, the number of permutations was set to 10,000. In order to determine which microbial taxa are primarily responsible for the differences between groups of samples, we performed partial least squares-discriminant analysis (PLS-DA), a supervised multivariate dimensionality reduction and classification technique using the R package “mixOmics” (Liquet et al., 2012; Rohart et al., 2017). Throughout our longitudinal analyses, the dependence on multiple samples per individual at different visits was considered. Therefore, to account for the clustering of samples by patients, we used the *strata* term in the *adonis* function to restrict the permutations within the samples from each patient for the PERMANOVA models. Additionally, we used multilevel partial least squares-discriminant analysis (mPLS-DA), which accounts for clustered samples (from the same patient) and correlations among microbial taxa.

After the PERMANOVA tests, non-parametric univariate tests were conducted at the taxonomic levels, where significant differences were observed to identify the taxa that contributed to those differences. We used Mann-Whitney-U-tests and Wilcoxon signed-rank tests for unpaired (patients vs. HCs) and paired (patients at different visits) respectively. The false discovery rate (fdr) approach was used to correct for multiple comparisons.

### Analysis clinical variables associated with microbiome composition

PERMANOVA was also performed to examine the factors associated with microbiome composition at the different visits. For each model, the effect of laxative use was corrected by including it as the first independent variable. At T0, the association between microbiome composition and weight loss, illness duration, BMI-SDS, Kcal, and leptin was tested. We also examined longitudinally the combined and individual effects of BMI-SDS and Kcal on the microbiome composition (genus-level) considering all visits together. Additionally, the combined effects of BMI-SDS, leptin, and Kcal together and the effect of leptin alone (unadjusted) on the microbiome composition were investigated longitudinally considering visits T0, T7, and T8.

### Analysis of prognostic relevance of baseline microbiome for clinical outcome

To investigate the association of the gut microbiota (genus- and ASV-levels) at admission (T0) or discharge (T7) with the different variables of clinical outcome (duration of treatment (only with admission microbiome), hospital readmission, and BMI-SDS at 1-year follow-up), first, a linear model (or logistic model for readmission) was constructed, where the effect of weight loss (calculated as the difference in BMI-SDS between disease onset (premorbid BMI-SDS) and admission to the clinic), duration of illness, and BMI-SDS at admission were regressed out to control for factors known to influence the duration of inpatient treatment. Then, the residuals of this model were used as the dependent variable in a second linear model, with microbial relative abundances as an independent variable while controlling for laxative use. For the prediction analyses, we used the square root of the transformed relative abundances of the core microbial taxa to account for non-normal distributions.

## Supporting information

all supplemental tables

## Acknowledgements

The work was partly funded by the ERA-NET Neuron “Microbiome Gut Brain Axis in Anorexia Nervosa (MiGBAN)” project sponsored by the European Union (EU) and the German Ministry for Education and Research (BMBF) under Grant 01EW1906A; the Deutsche Forschungsgemeinschaft (DFG) under the Grant SE 2787/3-1 (“The role of the intestinal microbiome regarding prognosis and therapy in adolescent Anorexia nervosa – clinical and translational analyses”); the Deutsche Forschungsgemeinschaft (DFG) Research Unit FOR5042 under Grant DE1614/4-1 (“The microbiome as a therapeutic target in inflammatory bowel diseases”); the Deutsche Forschungsgemeinschaft (DFG) Excellence Cluster under Grant EXC2167 (“Precision Medicine in Chronic 688 Inflammation PMI”).

The authors are grateful to Mrs. Katja Cloppenborg-Schmidt and Mrs. Yasmin Claußen for their excellent technical support, and to the Clinical Trial Center Aachen (CTC-A) for their support in conducting the study.

## Data availability statement

Data available on request due to privacy/ethical restrictions.

## Author contributions

JS, JB, and BHD designed the study; BD, HS, NM, VR, and JS recruited the participants and helped gather clinical data and stool and blood samples; HS and LK oversaw sample logistics. NA analyzed the stool samples; NA, AS, and AD performed statistical analyses. NA, AS, and JS wrote the first draft of the manuscript. All coauthors revised the manuscript and agreed to its publication.

---

**Fig S1:**
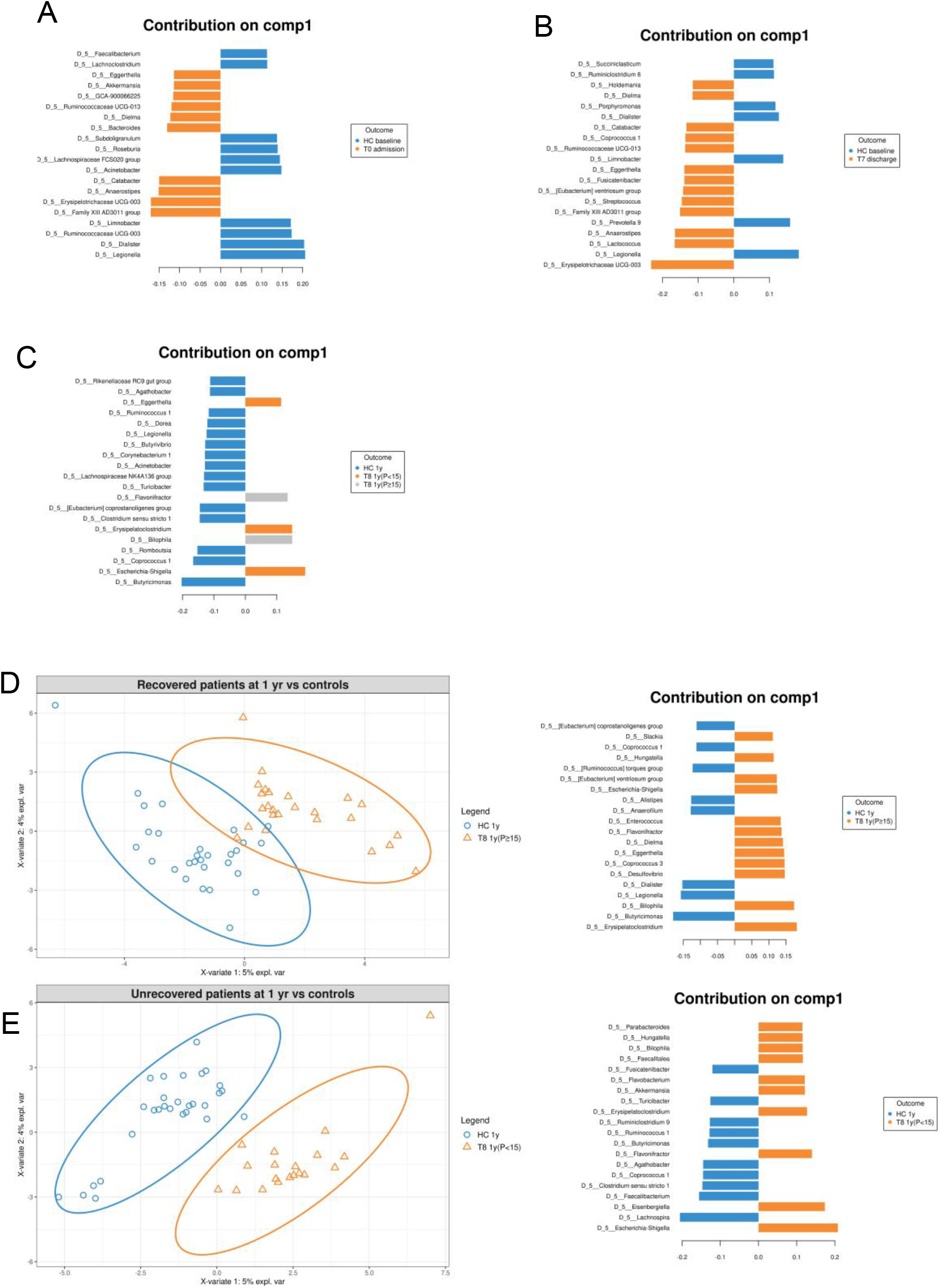
comparison of the microbiome of patients at admission/discharge and age-matched healthy controls. The figure reports the genera that mostly contribute to the discrimination of patients at admission **(A)** or discharge **(B)** and the HC group, and of patients at 1-year follow-up (divided into low-weight and recovered) and HCs **(C)**. Figure **(D)** shows the PLS-DA plot highlighting differences at the genus-level between recovered patients and the HC group. **(E)** The PLS-DA plot highlighting differences at the genus-level between low-weight patients and the HC group. The genera that mostly contributed to the discrimination are shown on the right side of each figure.

